# Personalized Virtual Reality Future Selves Elicit Introspective Brain Activation in Early Substance Use Disorder Recovery

**DOI:** 10.64898/2026.01.23.26344667

**Authors:** Brandon G Oberlin, Mario Dzemidzic, Yitong I Shen, Andrew J Nelson

## Abstract

Substance use disorder (SUD) recovery typically requires transformative change and prioritizing long-term healthy goals. Unfortunately, successful recovery is threatened by relapse rates that often exceed 50% in the first year. We previously reported on an experiential virtual reality (VR) SUD recovery intervention using personalized future self-avatars that produced emotional engagement and positive behavioral change, ie, stronger connection with the future self and future rewards and reduced craving. Here, we used fMRI to identify brain engagement to a future self experience with divergent futures.

Twenty adults (14 male, 33 years old) in early SUD recovery (<1 year) interacted with age-progressed versions of themselves in two different VR future ‘realities’: an SUD Future Self and a Recovery Future Self. Vivid lifelike visual and audio animation was augmented with a personalized narrative concerning future drug use and recovery. MRI immediately followed. Participants viewed videos of their future selves in the virtual environment and were directed to contemplate what they were seeing.

Viewing and contemplating the future selves elicited activation in midline default mode regions (posterior cingulate and ventromedial prefrontal cortices), visual regions including the occipital and fusiform face areas, and left middle frontal gyrus. The Recovery Future Self produced significant left occipital face area activation compared with the SUD Future Self. Midline default mode activation correlated with VR-induced increases in delayed reward preference, and also with greater trait perseverance.

Using digital selves as therapeutic agents reveals an entirely novel set of possible interventions and opens exciting new frontiers in behavior change methodology. Future studies targeting decision-making and future behavior could be informed by evaluating increased midline default mode engagement, with uniquely self-focused mechanisms signaled by executive network and face area coactivation. New hope for treatment-resistant mental health conditions is offered by the nearly limitless range of therapeutic experiences enabled by immersive digital therapeutics.

**Plain Language Summary:** High relapse rates in early recovery remains a serious challenge. To promote better outcomes, our team recently developed a virtual reality experience where people interacted with future versions of themselves. We used magnetic resonance imaging (MRI) to understand how the brain activated to this experience, and what brain responses were linked to positive outcomes.

We worked with 20 adults in early recovery. Each person used virtual reality to interact with two different future selves: one who had returned to substance use, and one who had stayed in recovery. These digital future selves looked and sounded like the participants and were paired with a personalized story about future drug use and recovery. Right after the virtual reality session, participants’ brains were scanned while they watched videos of these future selves and were asked to think about what they were seeing.

When people viewed and reflected on their future selves, brain areas involved in self-reflection and imagining the future became more active, along with regions that process faces. The future selves triggered brain activation in “self-focused” brain networks and in face-processing regions. Activity in key “self-focused” brain regions was linked to choosing larger, delayed rewards over smaller, immediate ones, and to lower impulsivity.

These findings suggest that lifelike digital versions of people’s future selves engage brain systems that support thinking ahead, persistence, and valuing long-term outcomes. This creates a promising new avenue for immersive digital therapeutic experiences to encourage lasting behavior change in early recovery from substance use disorder.

## Introduction

Substance use disorder (SUD)^1^ is widely regarded as a chronic condition (NIDA, 2020) requiring effective long-term aftercare (ASAM, 2014; Standard VI). Relapse occurs at high rates (50-90% within six months), even with evidence-based treatment (Weiss et al., 2011, Brecht and Herbeck, 2014, Brecht et al., 2005, Moeeni et al., 2016), highlighting the urgent need for better treatment outcomes. Strong recovery support greatly improves long-term outcomes, up to doubling SUD remission in longitudinal follow-up years later (Chi et al., 2011). Emergent technologies may improve SUD treatment aftercare, with virtual reality particularly attractive for its capacity to deliver highly salient personalized therapeutic experiences.

The human ability to organize behavior for optimized future outcomes relies in large part on the capacity for visualizing and identifying with possible futures and selves (Markus and Nurius, 1986, Bixter et al., 2020). When visualizing a personal future, those with SUD have shorter time horizons and less detailed imagined futures compared to controls (Manganiello, 1978, Petry et al., 1998, Mercuri et al., 2015). This impairment plausibly explains why those with SUD overly discount the value of delayed rewards (meta-analyses; Amlung et al., 2019, Amlung et al., 2017, MacKillop et al., 2011) and focus on the present rather than the future (Keough et al., 1999). Valuation of delayed rewards is directly linked to the degree of connection with the future self (Bartels and Rips, 2010, Ersner-Hershfield et al., 2009). Increasing the psychological connection to the future self with immersive digital simulations portraying future selves increases delayed reward preference (Hershfield et al., 2011, Shen et al., 2022) and healthier behaviors (Fox and Bailenson, 2009). Construal-level theory (Trope and Liberman, 2010, Mishra et al., 2020) predicts that making the abstract more concrete amplifies the salience of the future, and in the context of SUD remission, increases the attractiveness of recovery rewards and aversiveness of drug use. Therefore, immersive digital simulations using future selves could offer a practicable technique to promote adaptive, pro-recovery choices and healthier outcomes.

We created a highly personalized immersive virtual reality (VR) experience for participants in early SUD recovery (Shen et al., 2022). These vivid animated renderings of age-progressed selves narrated the events of two possible futures in their own voice. First traveling to the SUD Future, and then to the Recovery Future, participants experienced future selves who described profoundly divergent outcomes for next 15 years. Face, body, voice, and content were tailored to enhance vividness—an important factor to increase prosocial behavior change (Van Gelder et al., 2015). The spoken content was constructed from participants’ own stated costs and benefits, in the fashion of motivational interviewing (Bischof et al., 2021). This experiential therapy integrated future self-continuity (Hershfield, 2011, Hershfield et al., 2011) and self-discrepancy (Ogilvie, 1987, Paternoster and Bushway, 2009) to increase salience and the psychological connection with future selves. Permutations of the future self approach have been proposed for smoking (Şenel and Slater, 2020) and used to increase exercise (Fox and Bailenson, 2009). Our VR experience was iteratively designed together with members of the local SUD recovery community and specifically intended to be emotional and transformative— echoing epiphany experiences often reported by those in recovery (Galanter et al., 2012, Lawford, 2009). While this intervention produced significant pro-recovery behavioral and self-reported improvements in a pilot study, the brain mechanisms governing these are unknown.

Brain activity related to the concept of ‘self’ resides largely in cortical midline structures, especially the ventromedial prefrontal cortex (vmPFC) and posterior cingulate cortex (PCC) (for review; Northoff, 2017). These central connectivity hubs (Tomasi et al., 2014) comprise the medial components of the default mode network, a distributed resting-state network widely implicated in future-oriented and self-reflective thought, ie, prospection and introspection (Andrews-Hanna, 2012, Spreng and Grady, 2010). While initially described as deactivating during goal-directed tasks, considerable work now implicates the DMN, particularly in self-referential tasks such as autobiographical planning (Spreng et al., 2010, Spreng, 2012, Spreng et al., 2015) and imagining the future or past (Addis et al., 2009). Converging evidence suggests that the DMN integrates prior experiences, personal identity, simulations of the future, and action intent to create an internal narrative that guides behavior (for review see Menon, 2023).

To best understand brain activation during an immersive VR experience, functional magnetic resonance imaging (fMRI) data would ideally be acquired *during* the VR experience. However, immersive VR using a head-mounted display is not readily deployed in MRI scanners, due to metallic components, and the serious limitations of presence and agency (Makransky and Petersen, 2021) imposed by MRI-compliant immobility. However, we reasoned that a video from a recently experienced VR environment, containing VR avatars, would be expected to elicit brain activity representing activation to the experience itself. We previously reported that our novel VR future self experience increased delayed reward preference and future self-continuity, and decreased craving in early-recovering SUD participants (Shen et al., 2022). We hypothesize that viewing the future self during fMRI, immediately following an immersive VR experience with future selves, will increase activation in introspective and prospective brain regions, relative to the control condition. Specifically, we predict increased activation in posterior and anterior midline default mode regions, ie, the posterior cingulate cortex (PCC) and ventromedial prefrontal cortex (vmPFC) to the future self. We also hypothesize that this default mode activation will correlate with the recovery-related factors responding to the intervention— delayed reward preference, future self-continuity, and craving (Shen et al., 2022). We additionally performed exploratory correlations between brain responses and SUD-relevant traits and quality of life factors related to DSM-5 criteria for SUD diagnosis.

## Methods

### 1. Study Design Overview

Study procedures were as reported in Shen et al. (2022). Briefly, participants were phone screened, interviewed in-person, and scheduled for a study day—avatars were constructed in the intervening time. On the study day participants underwent VR habituation, personality/behavioral assessments (pre- and post-VR), the VR intervention, and fMRI. For 30 subsequent days participants received daily smartphone reminders of their experience, ‘mRecovery’, and then provided longitudinal 30-day follow-up data. Study elements are illustrated in Figure 1, with the senior (first) author pictured as a participant.

**Figure 1.**
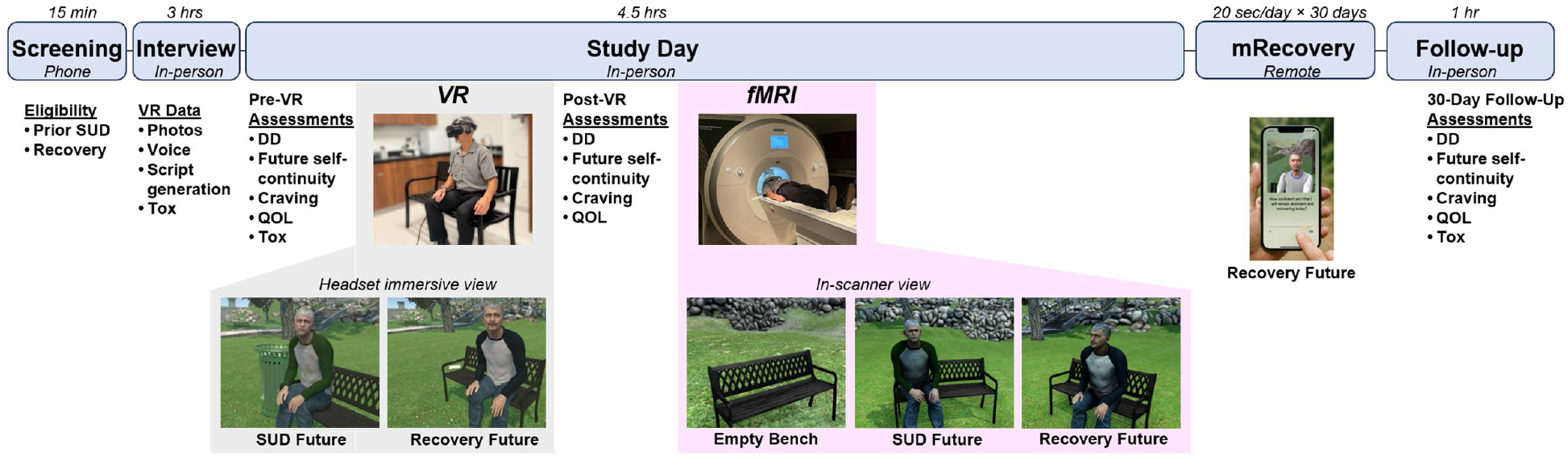
Study Procedures. (Screening) initial eligibility confirmed. (Interview) detailed characteristics collected, including data required for creating personalized VR experiences. (Study Day) Prior to, and following the VR experience, participants performed tasks and assessments. The VR experience (gray background) comprised a brief habituation, then interaction with future selves. Following assessments, participants were imaged on a 3T MRI scanner (pink background) while contemplating the virtual park and future selves shown. Views of videos displayed inside the VR headset and during MRI head coil are shown in the lower level. (mRecovery) daily images of the Recovery Future Self were sent to participants’ smartphones via MMS for 30 days. (Follow-up) Participants again completed assessments and self-reported drug/alcohol use. All in-person visits included toxicology screening (Tox); breath alcohol testing and urine screens. DD, delay discounting; QOL, quality of life survey. Approximate completion times shown above each study stage.

### 2. Participants

Twenty participants were recruited from treatment centers and recovery houses in and around Indianapolis, Indiana, targeting adults in recovery from alcohol and/or substance use disorders (SUD). Initial eligibility was determined via phone screening (including demographics, substance use history, treatment history, medical conditions, and current medications) to determine initial eligibility, which was confirmed by detailed in-person interviews (below). Study inclusion criteria were: between 21-50 years old, English fluency, diagnosis and treatment for use disorders of alcohol and/or illicit drugs, early SUD recovery (less than one year of continuous abstinence^2^), greater than 14 days abstinent from drug/alcohol use at the time of the study day (one participant with a single alcohol lapse six days prior to the study day was included), negative urine screen and breath alcohol results, and actively engaging in recovery activities. Comorbid psychiatric conditions were permitted to maximize generalizability. Exclusions were contraindications for VR or MRI, current use of alcohol or illicit drugs, >1 lapse event between the interview and the study day, not self-identified as being ‘in recovery’, disorders or history of neurological disease of cerebral origin, head injury with > 20 min loss of consciousness, or left-handedness. Participants signed written informed consent documents and were compensated ($250 total) for study completion, which included the 30-day follow-up appointment. All recruiting and study procedures were approved by the Indiana University Institutional Review Board (IRB Protocol #1805574553). *N*=21 were reported in Shen et al. (2022), but one of those participants was unable to complete MRI due to claustrophobia-related anxiety, resulting in the *n*=20 sample reported in this work. Two of these did not perform the delay discounting task due to Covid-19-related research restrictions. Participant characteristics are detailed in Table 1.

**Table 1.** Participant Characteristics (*N* = 20)

|  | Mean (SD) or n (%) |
| --- | --- |
| Age | 33.25 (7.50) |
| Sex (male) | 14 (70) |
| Race (white)<sup>a</sup> | 18 (90) |
| Years of education | 14.80 (3.12) |
| Days of abstinence | 73.80 (45.72) |
| Current nicotine use | 13 (65) |
| Family history ASUD<sup>b</sup> | 12 (60) |
| Recovery activities per month<sup>c</sup> | 24.30 (15.71) |

<sup>a</sup>Additionally, one black and one multiple-race participant in sample
<sup>b</sup>At least one 1<sup>st</sup> degree relative with alcohol and/or illicit substance use disorder (ASUD).
<sup>c</sup>Includes clinical appointments and group meetings.

### 3. Interview

The in-person interview comprised elaborated demographic information, nicotine use, family history of SUD, and quality of life survey. SUD symptom counts for alcohol and the three most-used substances were quantified with the structured clinical interview for DSM-5 disorders (SCID-5-RV; First et al., 2015). The Timeline Follow-back (TLFB; Sobell et al., 1986) characterized participants substance use patterns 35 days prior to treatment. Accuracy was enhanced with memorable event dates, past text messages, and any other useful memory triggers. Alcohol and illicit drug use was characterized by type and amount, and SUD family history was quantified by degree of relatedness with probable SUD positive relatives. We also characterized current recovery-related activities, treatment history, and confidence in remaining abstinent. Characteristic visual, audio, and content was collected for personalized avatar creation, detailed below.

### 4. Avatar and Script Generation

The avatars were constructed from studio-lit high-resolution photos (three face angles, two full-body angles, and close-ups of the eyes). Voice recording captured participants’ tonal quality, which was approximated with manual adjustment to Google’s text-to-speech engine. The phrasal template was personalized with information provided by the participant at the interview (names of loved ones, motivations for maintaining recovery, costs and punishments of prior drug use, and rewarding future goals, ie, “hopes and dreams”). The avatars were rendered three-dimensionally (Avatar SDK; https://avatarsdk.com/) with face aging features (custom texture and shader methods) and realistic eye movement and tracking (RandomEyes) implemented within an integrated VR environment (Unity 3D). The spoken script was synced with animated mouth and facial musculature. Some idiosyncratic features, primarily hairstyles and tattoos, required hand-editing to maximize personalization and realism. The two future selves were age-progressed to approximate 15 years into the future; this interval portrays sufficiently obvious visual markers of aging, and is intermediate to delays commonly used in temporal discounting tasks (eg, Madden et al., 1997).

### 5. Study day procedures

Urine screening (Wondfo Biotech, Ltd.) and breathalyzer confirmed ongoing drug and alcohol abstinence, and participants were queried for ongoing self-identification with ‘recovery’. Daily nicotine users were provided an appropriately dosed nicotine patch (CVS Pharmacy, Inc.) per manufacturer’s recommendations to prevent withdrawal (unless refused, *n*=2). Vital signs were taken to confirm healthy state, and lack of MRI contraindications was confirmed.

#### 5.1 Behavioral and Personality Assessments

##### Delay discounting

An adjusting-amount delay discounting (DD) task quantified relative preference for delayed rewards (Du et al., 2002, Oberlin et al., 2015). Choices between smaller immediate versus larger delayed monetary rewards adjusted the next trial’s immediate amount down or up, respectively, allowing the procedure to converge on indifference points. The $100 amount was delayed by 2 days, 1 week, 1 month, 6 months, 1 year, and 5 years in 30 choice trials (5 trials × 6 delays). Participants were instructed to choose according to their real preference, and that some choices would be selected at random and paid according to their choice. The actual payout was an additional $20 at the end of the study day, obfuscated by computer selection and rounding.

##### Future self-continuity

The validated future self-continuity scale indexes the perceived psychological distance to the imagined future self (Ersner-Hershfield et al., 2009). Participants endorsed future self-similarity and connectedness with Euler circle pairs labeled ‘Current Self’ and ‘Future Self’ that vary in degree of overlap. We used the original scale with two minor modifications: an additional option depicting complete overlap (Ersner-Hershfield et al., 2009, Shen et al., 2022); and an expanded temporal window of 15 years matching the age progression represented in the paradigm (the first four participants rated according to 10 years).

##### SUPPS-P Impulsive Behavior Scale

Derived from the original UPPS scale containing four subscales and 45 items (Whiteside and Lynam, 2001), this shortened version comprises 20 items across five subscales (negative urgency, lack of perseverance, lack of premeditation, sensation seeking, and positive urgency) is a reliable and valid low-burden alternative for assessing impulsivity traits (Cyders et al., 2014).

##### Zimbardo Time Perspective Inventory (ZTPI short form)

This shortened version of the original contains 22 items in two subscales that quantify time perspective, ie, orientation to the Future or Present (Keough et al., 1999).

##### Quality of Life (QOL)

The QOL survey (Flanagan, 1978) quantifies key aspects of subjective well-being that tend to improve in SUD remission (eg, Tracy et al., 2012). This 16-item scale is rated on a 7-point Likert-type scale ranging from “Delighted”(7) to “Terrible”(1) with increments labeled “Pleased”, “Mostly Satisfied”, “Mixed”, “Mostly Dissatisfied”, “Unhappy” in 16 items across 6 domains (well-being, relations with people, social/community activities, personal development, recreation, and independence).

##### VR questionnaire

Participants rated the VR experience on elements of presence (Sheridan, 1992), tolerability, cybersickness, and enjoyment. Items and scores were previously reported in (Shen et al., 2022; Table S2).

#### 5.2 VR Delivery

VR was delivered on a Samsung Odyssey (6 DOF, 90Hz, 2880×1600 pixels per-eye, 110° horizonal FoV) running on an Acer Predator laptop (Intel Core i7 CPU, Nvidia GeForce GTX 1060 GPU). VR experiences were conducted in a cleared laboratory space with a metal park bench, with the researcher closely supervising, managing the cables, and preventing stumbles or collisions. Interpupillary distance, headband tightness, and earphone placement was adjusted prior to beginning the experiences.

##### 5.2.1 VR Habituation

Participants experienced the virtual park, without avatars, for 4.62 ±1.79 minutes. The city park was nearly identical to the environment used for the intervention, complete with chirping birds, rustling of leaves in a breeze, and distant lawn mowing noise. Participants were instructed to sit on the park bench, which was spatially registered to a real metal park bench. This habituation session was designed to reduce novelty effects and familiarize participants to the environment where the key messaging would be delivered.

##### 5.2.2 VR Intervention

The VR paradigm featured personalized avatars representing the Present Self and age-progressed SUD Future Self, and Recovery Future Self. The intervention was designed to take ∼5 minutes to complete, although in practice took 5.12 ±0.53 minutes due to inter-subject variation. The intervention comprised four scenes: (1) white room introduction with Present Self, (2) SUD Future Self scenario, (3) Recovery Future Self scenario, and (4) debriefing in white room with Present Self. In scene 1 (white room setting), a disembodied narrator explained that this would be a simulated time travel experience 15 years into the future. Participants were instructed to turned around and view themselves in a mirror and move their head and hands. The mirrored Present Self matched participants’ movements via tracked head and hand positions and inverse kinematics solving for ∼30 seconds to establish ‘body transfer’ (Hershfield et al., 2011, Yee, 2007). The Present Self then appeared as a non-playable character and gestured to the participant to sit on the park bench. The Present Self referred to the participant, “I am you” and discussed personal details. The Present Self addressed the participant in the first scene using first person inclusive plural language (eg, “We enjoy…”, or “our main reason for…”), but upon advancing to the future—scenes 2 and 3—the future self used first person singular retrospective language. (eg, “I always thought…”, “Life turned out better than I imagined…”). The use of inclusive plural first person language was designed to increase connection with the future self by reducing psychological distance. Participants were then asked to “Choose your future” with two possible options. After choosing, the intervention transitioned into scene 2 (city park setting) where participants met their SUD Future Self. The SUD Future Self was unkempt, hunched forward, avoided eye contact, and discussed the past 15 years’ costs and punishments (from interview responses). The scene closed with the SUD Future Self saying, “don’t let this happen to us”. After a brief transportation back to the white room for a different choice, participants were transported to the city park to meet the Recovery Future Self in scene 3. The Recovery Future Self was poised and presented as healthy and positive, with a vague smile and steady eye contact. The Recovery Future Self described the past 15 years in terms of success and achievement of future goals (“hopes and dreams”). Finally, in scene 4, participants were transported back to the white room where they were debriefed by the Present Self. These closing comments highlighted the contingency between present action and future outcomes, promoted agency, and offered positive encouragement with confidence. The shirt colors were blue and green for SUD and Recovery futures selves for visual emphasis of different futures (randomized).

### 6. Neuroimaging

#### 6.1 MRI Acquisition

Imaging was conducted using a Siemens 3T Prisma MRI scanner (Erlangen, Germany) with a 32-channel head coil array. Structural imaging included T1-weighted MPRAGE (5 min 12 sec, 1.05×1.05×1.2 mm^3^ voxels) and was followed by a pair of 16-second phase-reversed spin echo scans to assess field distortion (5 A-P and 5 P-A phase direction volumes, TR/TE= 1370/51.60 ms, other parameters matched the BOLD scan parameters detailed below). The 13 min 51 sec virtual park video viewing scan used a multiband echo planar imaging (MB EPI) sequence from the Center for Magnetic Resonance Imaging at University of Minnesota (1018 blood oxygenation level dependent (BOLD) volumes, gradient echo, MB factor 4, repetition/echo time; TR/TE = 810/29 ms, 56° flip angle, 2.5 mm/side isotropic voxels, 220×220 mm^2^ field of view, 48 slices (Moeller et al., 2010)). Prior to the park video viewing task, participants completed a resting-state scan with eyes fixated on a white crosshair on the black background (10 min 7 sec, other parameters as in the park video scan). Following the virtual park videos, a delay discounting task and diffusion-weighted imaging were administered (diffusion-weighted imagining data will be reported elsewhere).

#### 6.2 Virtual Park Videos

Participants viewed videos of the three-dimensional park environment and avatars from the VR intervention. Three different videos (one with just the empty bench in the middle of the park, and two showing the animated future self avatars sitting on the bench in the park) were designed to engage the recent memory of the VR intervention. Each video was a single 3-minute camera shot comprised of a left to right dolly movement focusing on the avatar (or empty bench) from a viewpoint 2 meters away (constructed in Timeline and exported using Unity Recorder). The avatars were shown sitting on the bench as though waiting, using the same posture, dress, and mannerisms programmed for the intervention. Avatars did not interact with or acknowledge the viewer. This video presentation was intended to evoke the recent VR experience and engage introspection, positioning the viewer as an unseen observer. Participants were instructed to “Think about the place and person shown”. The videos were presented in the following fixed order: Empty Park Pre, SUD Future Self, Recovery Future Self, and Empty Park Post. To facilitate return to cognitive baseline, 30-second still images of the same park view were positioned after the Future Selves videos with 15-second stills positioned prior to and following the entire video block. In videos featuring the park bench and avatars, participants were instructed to button-press when a leaf fell behind the bench at fixed times (twice; 46 and 120 seconds from video start) to verify attentional engagement.

### 7. Neuroimaging Analyses

#### 7.1 Preprocessing

Image preprocessing used FEAT (FMRI Expert Analysis Tool) Version 6.00, part of the FMRIB Software Library (FSL; Jenkinson et al., 2012), including BOLD volume geometric distortion correction with *topup/applytopup* that utilized distortion field estimates from the spin echo field mapping scans, motion correction with *mcflirt*, brain extraction with *bet*, registration to each participant’s T1-weighted image and MNI152 standard space with *flirt* and *fnirt*, and 6mm FWHM Gaussian filter spatial smoothing. Subsequently, BOLD data were presented to FSL’s MELODIC version 3.15 to automatically estimate and retain an optimal number of independent components for each scan (Beckmann and Smith, 2004) and perform denoising with an unsupervised ICA-AROMA (Pruim et al., 2015) classifier. The resulting denoised data were assessed for residual head motion, transformed to a standard Montreal Neurological Institute (MNI) space, and interpolated to 2mm isotropic voxels for subsequent statistical analyses in SPM12 (Ashburner et al., 2014).

#### 7.2 Within-subject BOLD fMRI models

Within-subject fixed effects of the BOLD response to each of the conditions (eg, videos listed in 6.2) were estimated in SPM12 using the canonical hemodynamic response function (HRF). Six head motion parameters from realignment served as multiple regressors. The residual temporal autocorrelation was removed using SPM’s FAST approach appropriate for short TR data, while high-pass filter (1/128 Hz) removed low-frequency noise (Olszowy et al., 2019).

### 8. Mobile Health (mRecovery)

Daily texts displaying an image of the Recovery Future Self were sent to participants for 30 days. These were designed to elicit memory of the VR experience and invoke a positive future. Image viewing was verified with a single query (“how confident am I that I will remain abstinent today?”) that required a response on a 0-100% sliding scale (Qualtrics). The system was tested with participants on their smartphones just prior to discharge on the study day. Participants were familiarized with the system and informed of bonus incentives for compliance.

### 9. 30-Day Follow-Up Procedures

Longitudinal follow-up appointments included urine drug screen, breath alcohol testing, and self-reported drug use (positive responses triggered TLFB interview to quantify use amount, frequency, and type). Delay discounting, future self-continuity, and QOL were assessed again. All responses to the mRecovery reminders were logged. The actual follow-ups occurred at 37 ±8.8 days due to scheduling variability; *n*=2 completed follow-up over phone due to Covid-19-related disruptions.

### 10. Power and Statistical Analysis

#### 10.1 Planned Analyses

The study was powered for behavioral effects, with *N*=18 sufficient at power = 80% and alpha =.05 (G*Power 3.1.9.7) based estimates from prior work (Hershfield et al., 2011), as previously reported (Shen et al., 2022). Planned analyses included random effects models of brain responses to avatars, [Futures > Empty Park], and the inverse, [Empty Park > Futures]. The empty park control contained all visual and audio elements of the experience *except* the future self avatar. Significant brain responses to futures were tested for associations with effects (behavioral and self-report) previously reported in Shen et al., (2022), ie, increased delayed reward preference, increased future self-continuity, and decreased craving. These were Δ%DD AUC, Δfuture self-similarity, and Δfuture self-connectedness, and Δcraving for pre-VR versus study day and pre-VR versus follow-up; 8 factors total. Alpha was set to 0.05 for all analyses. Significant clusters (functional regions of interest; fROIs) were tested for correlation with SUD recovery-specific outcomes, with false discovery rate set to <5%, ie, Benjamini-Hochberg (1995); *q*<.05 satisfied by *p*<.00625 (8 tests) for each cluster. Means and standard deviations are reported as (*M* ±*SD*) in text and tables.

#### 10.2 Exploratory Analyses

Exploratory correlations were performed with 13 potentially informative factors, where 7 related to SUD traits, and 6 to QOL aspects reflecting DSM criteria expected to improve in recovery. SUD trait-related factors were: ZTPI Future, ZTPI Present, and 5 SUPPS subscales. Changes in QOL (delta between study day and 30 days later) were: Health, Family, Love/Romance, Work, Social life, and Recreation. Exploratory analyses tested for associations with brain activation in 13 factors total. Significant clusters were tested for correlation, with false discovery rate *q*<.05 satisfied by *p*<.00385 (13 tests) for each cluster in exploratory analyses.

#### 10.3 Statistical Models

This future visualization intervention was designed to increase delayed reward preference and future self-continuity by increasing focus on the future. We combined both futures (SUD and Recovery) together for analyses, as both represented plausible outcomes for people in early recovery. Both empty parks were combined for the control condition. A group-level random effects model was constructed from participant-level contrasts of virtual park videos to yield [Futures > Empty Park] and [Empty Park > Futures] contrasts in SPM12. Significant clusters were identified by meeting family-wise error correction *p*_*FWE*_ < 0.05 at a cluster-forming threshold of *p*_*uncorr*_ <.001 (corresponding cluster size *k* ≥ 108). To identify behavioral intervention responses correlating with brain activation, mean contrast values were extracted from each cluster (MarsBar toolbox; https://github.com/marsbar-toolbox/marsbar). Correlations between extracted values and key behavioral changes were tested in SPSS (v29; IBM). We performed exploratory correlations between 13 key SUD and QOL factors and both [Futures > Empty Park] and [Empty Park > Futures] contrasts. To examine if the two futures differed from each other, [SUD Future > Recovery Future] and [Recovery Future > SUD Future] were examined for differences within areas activated by any future thinking (ie, masked by [Futures > Empty Park]).

## Results

### 2. Neuroimaging

fMRI activation and correlations are reported below. Behavioral and self-reported data were previously reported in Shen (2022), including sustained drug/alcohol abstinence at 30 days for 18 out of 21 participants.

#### 1.1 Virtual Park Videos Whole-Brain Results

Videos of avatars sitting on the park bench (versus control) elicited activation primarily in midline and left lateral default mode network as well as visual attention areas. Seven significant clusters of activation in the [Futures > Empty Park] contrast were detected: posterior cingulate cortex, ventromedial prefrontal cortex (extending caudally into subgenual anterior cingulate), bilateral inferior occipital cortex (extending dorsally into left angular gyrus) that included occipital and fusiform face areas, left fusiform gyrus, left superior frontal gyrus, and left middle frontal gyrus, figure 2. The opposite contrast, [Empty Park > Futures], indicating deactivation, included bilateral lingual gyrus, supramarginal gyrus, precuneus, bilateral lateral temporal area (planum temporale), and right middle frontal gyrus; see table 2 for details.

**Table 2.** Significant Clusters: Viewing Virtual Future Selves and Park.

| *[Future Selves > Empty Park]* |  |  |  | Peak MNI Coordinates (mm) |  |  | Response to Future Selves<sup>1</sup> |
| --- | --- | --- | --- | --- | --- | --- | --- |
| Brain Region | *k* | Cluster *p*<sub>FWE</sub> | Peak voxel *Z* | *x* | *y* | *z* |  |
| R iOC | 2862 | <.001 | >8.00 | 40 | -82 | -16 | 0.45 ±0.24* |
| L iOC | 1912 | <.001 | 6.81 | -46 | -76 | 0 | 0.37 ±0.21* |
| L FuG | 141 | .015 | 4.70 | -36 | -52 | -20 | 0.28 ±0.35* |
| L SFG | 405 | <.001 | 4.48 | -12 | 60 | 22 | 0.25 ±0.23* |
| PCC | 560 | <.001 | 4.33 | -6 | -52 | 26 | 0.37 ±0.51* |
| vmPFC | 240 | .001 | 4.22 | -4 | 60 | -16 | 0.31 ±0.34* |
| L MFG | 137 | .018 | 4.16 | -46 | 12 | 44 | 0.26 ±0.33* |
| *[Empty Park > Future Selves]* |  |  |  |  |  |  |  |
| R LiG | 2397 | <.001 | 6.25 | 18 | -68 | -8 | -0.42 ±0.27* |
| L LiG | 1096 | <.001 | 5.84 | -24 | -48 | -10 | -0.42 ±0.32* |
| R SMG | 167 | .006 | 4.51 | 54 | -36 | 42 | -0.20 ±0.30* |
| R PCu | 310 | <.001 | 4.43 | 10 | -54 | 52 | -0.23 ±0.19* |
| R PT | 117 | .036 | 4.20 | 54 | -24 | 8 | -0.06 ±0.31 |
| R MFG | 217 | .001 | 4.11 | 36 | 8 | 46 | -0.18 ±0.19* |
| L PT | 171 | .006 | 4.00 | -44 | -26 | 2 | -0.02 ±0.37 |

<sup>1</sup>Mean ±SD contrast values within the cluster for the combined future selves condition, with significant mean activation (+) or deactivation (-) indicated by an asterisk (*p*<.05 in one-sample *t*-test versus zero). *k*=cluster size in voxels; MNI = Montreal Neurological Institute.
Anatomical regions: iOC, Inferior occipital cortex; FuG, Fusiform gyrus; SFG, Superior frontal gyrus; PCC, Posterior cingulate cortex; vmPFC, Ventromedial prefrontal cortex; MFG, Middle frontal gyrus; LiG, lingual gyrus; SMG, Supramarginal gyrus; PCu, Precuneus, PT, Planum temporale; R and L, right and left, respectively.

**Figure 2.**
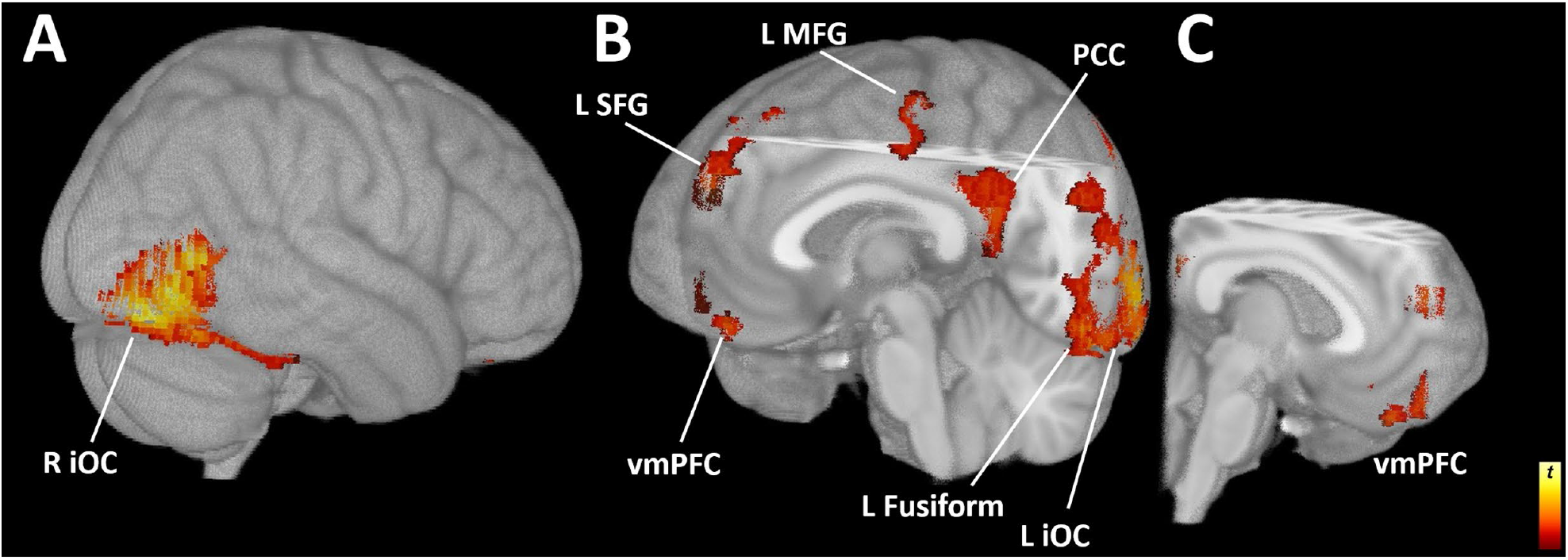
Activation During Future Self Contemplation. Activation in the [Futures > Empty Park] contrast produced 7 significant clusters. (A) View from rear right illustrates the right inferior occipital cortex (R iOC); (B) the cutaway view from the left front shows the ventromedial prefrontal cortex (vmPFC), left superior frontal gyrus (L SFG), left middle frontal gyrus (L MFG), the posterior cingulate cortex (PCC), and left inferior occipital cortex (L iOC); (C) the cutaway complement further illustrates vmPFC, PCC, and SFG; displayed at *p*_uncorr_<.001, *k>*100.

#### 1.2 Activation Correlated with Recovery-Relevant Responses

Eight behavioral and psychological responses to the VR intervention (Shen et al., 2022) were tested for correlation with mean contrast values from significant clusters from both [Futures > Empty Park] and [Empty Park > Futures]. The percent change in DD from pre-VR to the 30-day follow-up was correlated with activation to the future selves, [Futures > Empty Park], in the posterior cingulate cortex *r*(16)=.644, *p*=.004, *q*<.05 and ventromedial prefrontal cortex *r*(16)=.621, *p*=.006, *q*<.05; figure 3. No other significant results were detected in [Futures > Empty Park] contrast, and no significant correlations were detected in any regions from the [Empty Park > Futures] contrast.

**Figure 3.**
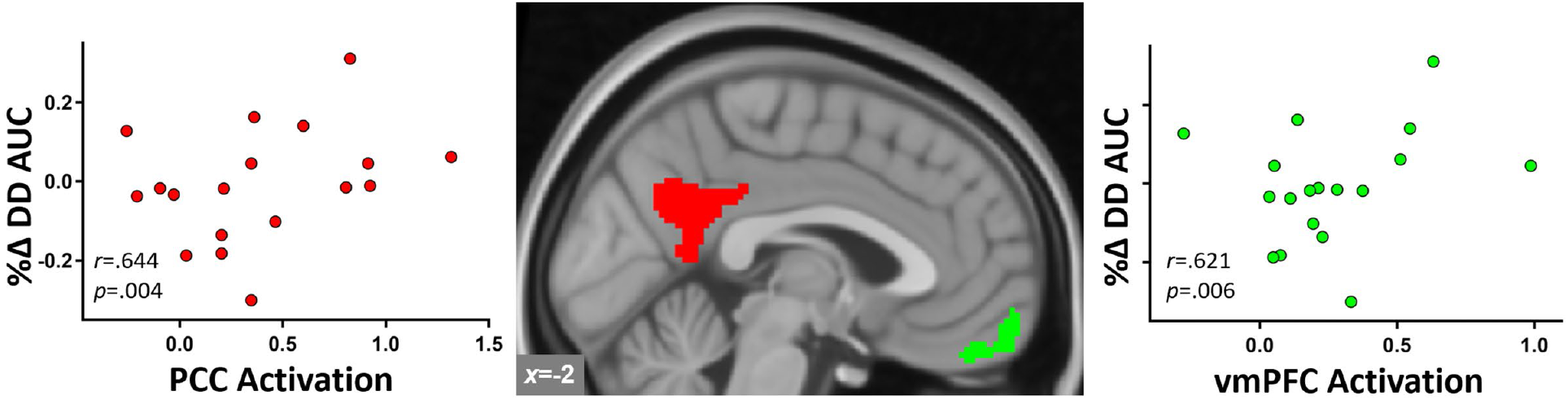
Brain Responses to Futures Correlated with Increased Delayed Reward Preference. [Futures > Empty Park] activation (mean contrast values) in posterior and anterior medial default mode regions (*center*). Activation in the (*left*) posterior cingulate (red), and (*right*) ventromedial prefrontal cortex (green) correlated with changes in delay discounting (AUC; area under the curve) from pre-VR to the 30-day follow-up, *q*s<.05.

#### 1.3 Exploratory Correlations with SUD Traits and QOL Changes

Activation in the posterior cingulate cortex cluster correlated with greater perserverence; that is, negatively with UPPPS Lack of Perseverance *r*(18)=−.636, *p*=.003, *q*<.05; figure 4, as measured in the inventory. No other significant results were detected in [Futures > Empty Park] contrast, and no significant correlations were detected in any regions from the [Empty Park > Futures] contrast.

**Figure 4.**
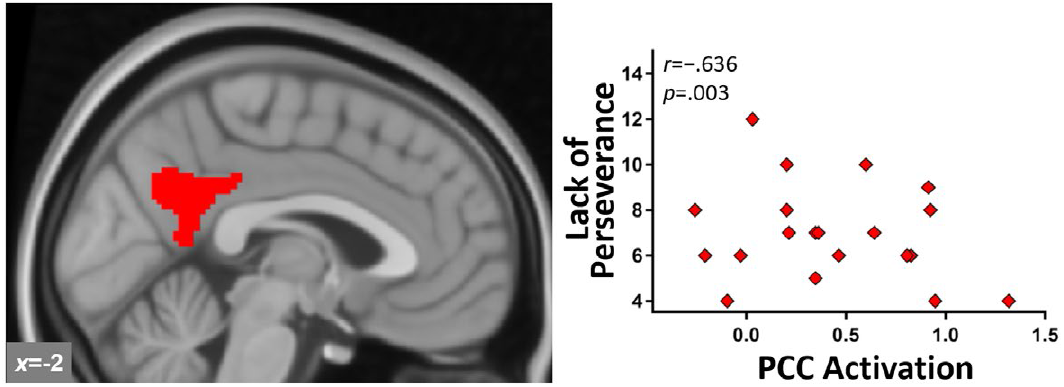
Posterior Cingulate Activation and Impulsivity. [Futures > Empty Park] activation in posterior cingulate cortex (*left*) negatively correlated with the UPPS Lack of Perseverance trait impulsivity measure (*right*), *q*<.05.

#### 1.4. Recovery and SUD Futures

Significant activation was detected in the [Recovery Future > SUD Future] in the inferior occipital cortex (occipital face area) figure 5, but no activation was detected in the opposite contrast, [SUD Future > Recovery Future].

**Figure 5.**
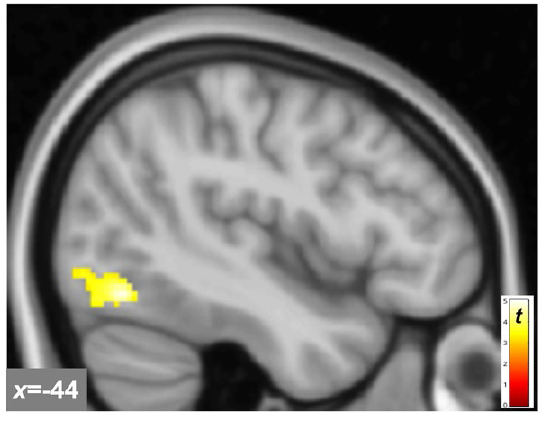
Brain Activation to Recovery Future. [Recovery Future > SUD Future] elicited significant activation in the occipital face area of the left inferior occipital cortex.

## Discussion

We previously demonstrated that interactions with age-progressed selves in an SUD or recovery future increased connection with the future self, increased valuation of future rewards, and reduced craving (Shen et al., 2022). To ascertain what brain systems were engaged by the VR experience, we presented a video of the future selves in the same virtual park during fMRI. This was designed to evoke the same prospective and introspective processing that occurred during the immersive VR experience. Brain activation to the future selves was largely concentrated in default mode and visual attention regions. The activation to future selves in midline default mode (PCC and vmPFC) correlated with increased valuation of delayed rewards. Further, PCC activation to the future selves correlated with greater perseverance (trait impulsivity subscale). While activation between the SUD Future Self and Recovery Future Self mostly overlapped, greater activation to the Recovery Future Self was detected in the inferior occipital cortex, indicating engagement of visual attention and associative processes specific to the recovery future. These findings suggest that this novel intervention recruits default mode network activity, and that this inwardly focused cognition shifts behavioral preference toward the future.

The visual scene presented during fMRI was designed to project attention into imagined and “pre-experienced” futures, ie, by simultaneously eliciting autobiographical mental time travel, perspective-taking, introspection, and prospection. The intervention engages four major modes of prospection: simulation, prediction, intention, and planning (Szpunar et al., 2014). A large body of work supports the central role of the DMN in these internally-generated processes, ie, stimulus-independent thought (Spreng and Grady, 2010, Menon, 2023, Andrews-Hanna, 2012, Andrews-Hanna et al., 2014). In our study, these internal states are induced by visual stimuli, which include the age-progressed self (future self avatars) and the environmental context representing the non-self future (city park), ie, other aspects of the future minus the future self. Tasks such as mental time travel, mental simulation, and scene construction recruit DMN regions in these prospective and introspective processes (Schacter et al., 2007, Cabeza et al., 1997, Foster et al., 2012, Addis et al., 2007), with particular focus on the self (meta-analysis; Qin and Northoff, 2011). Activation to future selves in the current study reflects activity in the central hubs of the DMN; the PCC and vmPFC (Yeo et al., 2011, Tomasi et al., 2014) supporting the DMN’s role in animating a deep internal narrative. While cognitive processes engaged by ‘thinking about what you see’ (instructions provided to participants for viewing the empty park bench or future selves during MRI) are technically unknowable, we infer that participants contemplated their personal futures, as they were immersed in an experience with future selves in the same visual environment just an hour earlier.

The VR intervention is designed to induce mental projection into the future that is both semantic (general) and episodic (specific). That is, participants interact with two semantic future selves that have lived according to divergent general strategies—one continuing to misuse drugs, and the other engaged in recovery. The episodic elements narrated by the future selves are events such as drunk driving accident (SUD future) or becoming a homeowner (recovery future). The park + avatar video during fMRI evokes the VR future simulation (albeit without avatar interaction) in a manner designed to induce contemplation. The semantic aspects are presumably linked to mind wandering, and therefore DMN activation, but the combined episodic elements might be expected to engage brain areas linked to intention and planning, ie, frontoparietal network. Converging evidence suggests that when planning is mixed with introspection (as in the current study), default mode systems co-activate with networks mediating goal-directed behavior; particularly the frontoparietal network. Prior work demonstrated that autobiographical planning produced correlated DMN and frontoparietal network activity, whereas a visuospatial planning task did not (Spreng et al., 2010). Other research examining the preparatory phase of verbal working memory execution indicates DMN coactivation with frontoparietal and inferior occipital cortex, ie, visual attention and association networks (Koshino et al., 2014). Consistent with executive engagement, our findings in the left MFG (dorsal aspect) and left iOC (angular gyrus extent) overlap with the frontoparietal network (Yeo et al., 2011), demonstrating at least left-lateralized frontoparietal coactivation. Contemplating a primed personal future after an immersive VR experience (containing both semantic and episodic elements) activates introspective default mode and executive frontoparietal network regions simultaneously.

The degree of midline default mode activation to future selves appeared to index capacity for increasing delayed reward preference. Discounting of delayed rewards decreased after the intervention, suggesting that the future self VR experience shifted attention and preference toward future rewards. Prior work indicates that the key regions responsible for performing delay discounting tasks are largely localized to the default mode, frontoparietal, and salience (resting state-defined) networks (McClure et al., 2004, Kable and Glimcher, 2007, Kable and Glimcher, 2010, Wesley and Bickel, 2014 [meta-analysis], Moro et al., 2023), which are implicated in psychopathology more generally (the ‘triple network model’; Menon, 2011). While delay discounting is a complex decision task^3^ that recruits various brain regions to execute, the brain regions that respond specifically to *interventions* that increase delayed reward preference fall largely within the midline default mode regions PCC and vmPFC (Oberlin et al., 2020, Stawarczyk and D’Argembeau, 2015). Moreover, the self-referential ‘cortical midline system’, which is deeply involved with the self, familiarity, and making self/other distinctions (Qin and Northoff, 2011, Musholt, 2013, Northoff, 2017, Andrews-Hanna et al., 2014) comprises the midline components of the default mode network. In the current work, PCC activation to the future selves correlated with lower trait impulsivity (lack of perseveration), suggesting a possible trait-level influence. The physical overlap and functional relationship between the PCC and vmPFC with self-focused thinking, prospection, and augmented delayed reward preference advances a mechanistic explanation: making the future self more real and ‘tangible’ increases prospective brain activation and shifts preference towards future rewards.

Activation specifically to the Recovery Future Self, relative to the SUD Future Self, was localized in the occipital face area, outside the default mode network. The occipital face area integrates visual face perception (Haxby et al., 2000) with episodic and semantic information (Ambrus et al., 2019, Eick et al., 2020) as part of the larger visual ventral stream (a brain system that derives object identity from raw visual information (Rust and DiCarlo, 2010, Ritchie et al., 2024)). In concert with the adjacent fusiform face area and lateral occipital cortex, the occipital face area comprises key nodes in the core early face-processing network (Nagy et al., 2012, Gschwind et al., 2012). Beyond semantic information linked to face identity, this region also responds to episodic recall (Addis et al., 2009). Further, functional connectivity with this location and lateral prefrontal cortex in episodic simulation (Demblon et al., 2016) or with joint default mode and frontoparietal activation to mental simulation (Gerlach et al., 2011) indicates its role in visual scene construction. Given that the Recovery Future Self delivered specific verbal messaging about a particular future, this information is presumably encoded with the visual information, linking this particular face to a panoply of semantic details about an imagined future—one in which recovery from addiction has provided considerable benefit compared with the alternative.

Identifying key brain regions to mark successful SUD recovery is not as straightforward as might be presumed. Most prior work attempting to answer this question used drug cue reactivity paradigms to mark success or failure; however, the utility of this approach may be limited to identifying regions rather than clear directional interpretations. That is, brain activation to drug cues may reflect either drug reward drive (implying greater risk) *or* active resistance to temptation, ie, recovery reward drive and abstinence-related payoffs (suggesting lower risk).

One or the other interpretation may be favored by particular paradigms or even vary between individuals. Nonetheless, considerable prior work implicates midline default regions PCC and vmPFC in predicting abstinence in early recovery. PCC activation to visual cocaine cues correlated with worse treatment outcomes and differentiated abstainers from relapsers (Kosten et al., 2006). During an executive functioning task, PCC activation was a predictor of relapse in the comparison of smoking satiety versus short-term abstinence (Loughead et al., 2015). As the PCC is a structural and functional rich club (Lord et al., 2017), its central role in connectivity might be the most informative. Broadly, greater PCC connectivity with other networks and within the default mode network tends to be associated with better SUD treatment outcomes (see Wilcox et al., 2019 for review). Activation in the anterior aspect of the midline default mode network, ie, the vmPFC, predicts abstinence-related outcomes, but with mixed directionality. Using visual alcohol cues, activation in the vmPFC correlated with craving and time to first relapse (Bach et al., 2015), whereas greater activation to neutral relaxing cues predicted relapse to alcohol (Seo et al., 2013, Blaine et al., 2017). However, greater vmPFC responsivity to visual heroin cues predicted better treatment adherence (Wang et al., 2015). Beyond the default mode network, left-lateralized frontoparietal (EEG) connectivity between the dlPFC and posterior parietal cortex differentiated early recovering AUD sustaining abstinence versus those who relapsed (Januszko et al., 2021). This pattern of frontoparietal connectivity resembles the left-lateralized frontoparietal and midline default mode coactivation observed in the present report. Together, the regions and networks showing activation and behavioral correlation reported here map onto key regions that predict relapse and likely underly recovery-related brain function.

## Limitations

Some limitations should be acknowledged. Significant findings in this modest sample inspired confidence in the effects (Lenth, 2001) and informed the approach (a larger ongoing RCT based on these methods); nonetheless, results in a larger sample would enhance generalizability. We detected main effects and correlations of activations to the future self relative to the control condition in early recovery SUD, but we cannot know how this might extrapolate to healthy controls. Main effects of activation to a personal future were identified, insofar as the virtual park video viewing reflected brain activation to the VR experience. As fMRI was performed within an hour of the VR experience, and the park viewing was a video of the same 3D environment (and avatars), this is presumably a reasonable assumption. While one might speculate that the ideal control condition for detecting activation to a future self would be the Present Self, recall that the VR experience never showed the Present Self in the virtual park. That is, the Present Self was never associated with the virtual park. Thus, to best capture the effect of the future selves—the putative active ingredient of this experiential therapy—the proper control is the park without either future self in the visual field. Another limitation is the use of a fixed order during imaging, which potentially limited the capacity to detect differences between the two futures. The fixed order reflected the VR presentation order, which was fixed for clinical reasons (recency effect favoring the optimistic future was designed to lower the psychological danger potentially caused by the SUD future self). A larger sample with only one future self in a between-subjects design could resolve this issue. VR development necessarily limited subject numbers, precluding this approach.

## Conclusion

VR permits bringing experiential therapies to life and implementing them in the clinic. We demonstrated that an autobiographical future simulation elicits positive pro-recovery effects (reduced impulsive choice and craving and increased identification with the personal future, along with low rates of relapse). This intervention produces semantic (introspective, contemplative) midline DMN and episodic (prospective, executive planning) frontoparietal coactivation while evoking general and specific SUD recovery elements. Our current report on brain engagement suggests that introspective and executive brain networks are involved with this reoriented future-focus, which corresponds with positive behavioral changes. The nascent digital therapeutic space is replete with new possibilities, allowing emerging technology to augment existing evidence-based techniques.

## Data Availability

All data produced in the present study are available upon reasonable request to the authors

## Acknowledgements

We are grateful to Dr. Elizabeth Lungwitz, research MRI personnel of the Research Imaging Core in the Medical Imaging Research Institute, Department of Radiology and Imaging Sciences, Dr. Yu-Chien Wu, Michele Dragoo, Traci Day, and Robert Bryant for excellent technical assistance, and for the generous support of the Indiana Clinical and Translational Sciences Institute Project Development Team (PRMC UL1TR001108) and the Indiana University Department of Psychiatry. We additionally thank the anonymous reviewers and editors for their insightful comments.

## Footnotes

1 For the purposes of this study, ‘SUD’ refers to alcohol and illicit drug use disorders (excluding nicotine and caffeine), reflecting the concept and definition of ‘addiction’ familiar to our research participants. While the DSM-5 recognizes nicotine and caffeine use disorders as SUD, these drugs are viewed differently, and their use is often tolerated or accepted by treatment programs and clients.

2 We defaulted to the definition commonly accepted among our participants and in our partner treatment centers. Out of 18 participants queried on their personal definition of recovery-compliant drug use, 15 participants endorsed complete abstinence from alcohol and all illicit drugs, and three participants allowed for some future use under specific conditions: variously, use of marijuana if legalized, occasional drinking, and hallucinogens following one year of total abstinence.

3 Considering delay discounting as the competition between reward drive state (valuation of specific reward properties and the degree of motivation) and delay penalty (optimism for the future and estimating probability of reward receipt) requires autonoetic future thinking, working memory, and executive inhibitory control, in addition to variable idiosyncratic functions, and obviously visual and motor systems.

## References

Addis, D. R., Pan, L., Vu, M.-A., Laiser, N. & Schacter, D. L. 2009. Constructive episodic simulation of the future and the past: Distinct subsystems of a core brain network mediate imagining and remembering. Neuropsychologia, 47, 2222–2238.

Addis, D. R., Wong, A. T. & Schacter, D. L. 2007. Remembering the past and imagining the future: Common and distinct neural substrates during event construction and elaboration. Neuropsychologia, 45, 1363–1377.

Ambrus, G. G., Amado, C., Krohn, L. & KovÁCs, G. 2019. TMS of the occipital face area modulates cross-domain identity priming. Brain Structure and Function, 224, 149–157.

Amlung, M., Marsden, E., Holshausen, K., Morris, V., Patel, H., Vedelago, L., Naish, K. R., Reed, D. D. & Mccabe, R. E. 2019. Delay Discounting as a Transdiagnostic Process in Psychiatric Disorders: A Meta-analysis. JAMA Psychiatry.

Amlung, M., Vedelago, L., Acker, J., Balodis, I. & Mackillop, J. 2017. Steep delay discounting and addictive behavior: a meta-analysis of continuous associations. Addiction, 112, 51–62.

Andrews-Hanna, J. R. 2012. The brain’s default network and its adaptive role in internal mentation. The Neuroscientist, 18, 251–270.

Andrews-Hanna, J. R., Smallwood, J. & Spreng, R. N. 2014. The default network and self-generated thought: Component processes, dynamic control, and clinical relevance. Annals of the new York Academy of Sciences, 1316, 29–52.

ASAM 2014. The ASAM Standards of Care for the Addiction Specialist Physician. American Society of Addiction Medicine.

Ashburner, J., Barnes, G., Chen, C.-C., Daunizeau, J., Flandin, G., Friston, K., Kiebel, S., Kilner, J., Litvak, V. & Moran, R. 2014. SPM12 manual. Wellcome Trust Centre for Neuroimaging, London, UK, 2464, 53.

Bach, P., Kirsch, M., Hoffmann, S., Jorde, A., Mann, K., Frank, J., Charlet, K., Beck, A., Heinz, A. & Walter, H. 2015. The effects of single nucleotide polymorphisms in glutamatergic neurotransmission genes on neural response to alcohol cues and craving. Addiction biology, 20, 1022–1032.

Bartels, D. M. & Rips, L. J. 2010. Psychological connectedness and intertemporal choice. Journal of Experimental Psychology: General, 139, 49.

Beckmann, C. F. & Smith, S. M. 2004. Probabilistic independent component analysis for functional magnetic resonance imaging. IEEE transactions on medical imaging, 23, 137–152.

Benjamini, Y. & Hochberg, Y. 1995. Controlling the false discovery rate: a practical and powerful approach to multiple testing. Journal of the Royal statistical society: series B (Methodological), 57, 289–300.

Bischof, G., Bischof, A. & Rumpf, H.-J. 2021. Motivational interviewing: an evidence-based approach for use in medical practice. Deutsches Ärzteblatt International, 118, 109.

Bixter, M. T., Mcmichael, S. L., Bunker, C. J., Adelman, R. M., Okun, M. A., Grimm, K. J., Graudejus, O. & Kwan, V. S. 2020. A test of a triadic conceptualization of future self-identification. PloS one, 15, e0242504.

Blaine, S. K., Seo, D. & Sinha, R. 2017. Peripheral and prefrontal stress system markers and risk of relapse in alcoholism. Addiction biology, 22, 468–478.

Brecht, M.-L., Anglin, M. D. & Dylan, M. 2005. Coerced treatment for methamphetamine abuse: Differential patient characteristics and outcomes. The American journal of drug and alcohol abuse, 31, 337–356.

Brecht, M.-L. & Herbeck, D. 2014. Time to relapse following treatment for methamphetamine use: a long-term perspective on patterns and predictors. Drug and alcohol dependence, 139, 18–25.

Cabeza, R., Mangels, J., Nyberg, L., Habib, R., Houle, S., Mcintosh, A. R. & Tulving, E. 1997. Brain regions differentially involved in remembering what and when: a PET study. Neuron, 19, 863–870.

Chi, F. W., Parthasarathy, S., Mertens, J. R. & Weisner, C. M. 2011. Continuing care and long-term substance use outcomes in managed care: early evidence for a primary care-based model. Psychiatric Services, 62, 1194–1200.

Cyders, M. A., Littlefield, A. K., Coffey, S. & Karyadi, K. A. 2014. Examination of a short English version of the UPPS-P Impulsive Behavior Scale. Addict Behav, 39, 1372–6.

Demblon, J., Bahri, M. A. & D’argembeau, A. 2016. Neural correlates of event clusters in past and future thoughts: How the brain integrates specific episodes with autobiographical knowledge. NeuroImage, 127, 257–266.

Du, W., Green, L. & Myerson, J. 2002. Cross-cultural comparisons of discounting delayed and probabilistic rewards. The Psychological Record, 52, 479–92.

Eick, C. M., KovÁcs, G., Rostalski, S.-M., RÖhrig, L. & Ambrus, G. G. 2020. The occipital face area is causally involved in identity-related visual-semantic associations. Brain Structure and Function, 225, 1483–1493.

Ersner-Hershfield, H., Garton, M. T., Ballard, K., Samanez-Larkin, G. R. & Knutson, B. 2009. Don’t stop thinking about tomorrow: Individual differences in future self-continuity account for saving. Judgm Decis Mak, 4, 280–286.

First, M. B., Williams, J. B. W., Karg, R. S. & Spitzer, R. L. 2015. SCID-5 for DSM-5, research version; SCID-5-RV, Arlington, VA, American Psychiatric Association.

Flanagan, J. C. 1978. A research approach to improving our quality of life. American psychologist, 33, 138.

Foster, B. L., Dastjerdi, M. & Parvizi, J. 2012. Neural populations in human posteromedial cortex display opposing responses during memory and numerical processing. Proceedings of the National Academy of Sciences, 109, 15514–15519.

Fox, J. & Bailenson, J. N. 2009. Virtual Self-Modeling: The Effects of Vicarious Reinforcement and Identification on Exercise Behaviors. Media Psychology, 12, 1–25.

Galanter, M., Dermatis, H. & Santucci, C. 2012. Young people in Alcoholics Anonymous: The role of spiritual orientation and AA member affiliation. Journal of addictive diseases, 31, 173–182.

Gerlach, K. D., Spreng, R. N., Gilmore, A. W. & Schacter, D. L. 2011. Solving future problems: default network and executive activity associated with goal-directed mental simulations. Neuroimage, 55, 1816–1824.

Gschwind, M., Pourtois, G., Schwartz, S., Van De Ville, D. & Vuilleumier, P. 2012. White-matter connectivity between face-responsive regions in the human brain. Cerebral cortex, 22, 1564–1576.

Haxby, J. V., Hoffman, E. A. & Gobbini, M. I. 2000. The distributed human neural system for face perception. Trends in cognitive sciences, 4, 223–233.

Hershfield, H. E. 2011. Future self-continuity: How conceptions of the future self transform intertemporal choice. Annals of the New York Academy of Sciences, 1235, 30.

Hershfield, H. E., Goldstein, D. G., Sharpe, W. F., Fox, J., Yeykelis, L., Carstensen, L. L. & Bailenson, J. N. 2011. Increasing Saving Behavior through Age-Progressed Renderings of the Future Self. J Mark Res, 48, S23–S37.

Januszko, P., Gmaj, B., Piotrowski, T., Kopera, M., Klimkiewicz, A., Wnorowska, A., WoŁyŃczyk-Gmaj, D., Brower, K. J., Wojnar, M. & Jakubczyk, A. 2021. Delta resting-state functional connectivity in the cognitive control network as a prognostic factor for maintaining abstinence: An eLORETA preliminary study. Drug and Alcohol Dependence, 218, 108393.

Jenkinson, M., Beckmann, C. F., Behrens, T. E., Woolrich, M. W. & Smith, S. M. 2012. Fsl. Neuroimage, 62, 782–90.

Kable, J. W. & Glimcher, P. W. 2007. The neural correlates of subjective value during intertemporal choice. Nat Neurosci, 10, 1625–33.

Kable, J. W. & Glimcher, P. W. 2010. An “as soon as possible” effect in human intertemporal decision making: behavioral evidence and neural mechanisms. J Neurophysiol, 103, 2513–31.

Keough, K. A., Zimbardo, P. G. & Boyd, J. N. 1999. Who’s smoking, drinking, and using drugs? Time perspective as a predictor of substance use. Basic and applied social psychology, 21, 149–164.

Koshino, H., Minamoto, T., Yaoi, K., Osaka, M. & Osaka, N. 2014. Coactivation of the default mode network regions and working memory network regions during task preparation. Scientific reports, 4, 5954.

Kosten, T. R., Scanley, B. E., Tucker, K. A., Oliveto, A., Prince, C., Sinha, R., Potenza, M. N., Skudlarski, P. & Wexler, B. E. 2006. Cue-induced brain activity changes and relapse in cocaine-dependent patients. Neuropsychopharmacology, 31, 644–650.

Lawford, C. K. 2009. Moments of clarity: voices from the front lines of addiction and recovery, Harper Collins.

Lenth, R. V. 2001. Some practical guidelines for effective sample size determination. The american statistician, 55, 187–193.

Lord, A. R., Li, M., Demenescu, L. R., Van Den Meer, J., Borchardt, V., Krause, A. L., Heinze, H.-J., Breakspear, M. & Walter, M. 2017. Richness in functional connectivity depends on the neuronal integrity within the posterior cingulate cortex. Frontiers in Neuroscience, 11, 184.

Loughead, J., Wileyto, E. P., Ruparel, K., Falcone, M., Hopson, R., Gur, R. & Lerman, C. 2015. Working memory-related neural activity predicts future smoking relapse. Neuropsychopharmacology, 40, 1311–1320.

Mackillop, J., Amlung, M. T., Few, L. R., Ray, L. A., Sweet, L. H. & Munafo, M. R. 2011. Delayed reward discounting and addictive behavior: a meta-analysis. Psychopharmacology (Berl), 216, 305–21.

Madden, G. J., Petry, N. M., Badger, G. J. & Bickel, W. K. 1997. Impulsive and self-control choices in opioid-dependent patients and non-drug-using control participants: drug and monetary rewards. Exp Clin Psychopharmacol, 5, 256–62.

Makransky, G. & Petersen, G. B. 2021. The cognitive affective model of immersive learning (CAMIL): A theoretical research-based model of learning in immersive virtual reality. Educational psychology review, 33, 937–958.

Manganiello, J. A. 1978. Opiate addiction: a study identifying three systematically related psychological correlates. Int J Addict, 13, 839–47.

Markus, H. & Nurius, P. 1986. Possible selves. American psychologist, 41, 954.

Mcclure, S. M., Laibson, D. I., Loewenstein, G. & Cohen, J. D. 2004. Separate neural systems value immediate and delayed monetary rewards. Science, 306, 503–7.

Menon, V. 2011. Large-scale brain networks and psychopathology: a unifying triple network model. Trends in cognitive sciences, 15, 483–506.

Menon, V. 2023. 20 years of the default mode network: A review and synthesis. Neuron, 111, 2469–2487.

Mercuri, K., Terrett, G., Henry, J. D., Bailey, P. E., Curran, H. V. & Rendell, P. G. 2015. Episodic foresight deficits in long-term opiate users. Psychopharmacology (Berl), 232, 1337–45.

Mishra, A. N., Raj, A. & Pani, A. K. 2020. Construal level research in decision making: Analysis and pushing forward the debate using bibliometric review and thematic analysis. Mishra, Arindra N, 106–135.

Moeeni, M., Razaghi, E. M., Ponnet, K., Torabi, F., Shafiee, S. A. & Pashaei, T. 2016. Predictors of time to relapse in amphetamine-type substance users in the matrix treatment program in Iran: a Cox proportional hazard model application. BMC psychiatry, 16, 265.

Moeller, S., Yacoub, E., Olman, C. A., Auerbach, E., Strupp, J., Harel, N. & UĞurbil, K. 2010. Multiband multislice GE-EPI at 7 tesla, with 16-fold acceleration using partial parallel imaging with application to high spatial and temporal whole-brain fMRI. Magnetic resonance in medicine, 63, 1144–1153.

Moro, A. S., Saccenti, D., Ferro, M., Scaini, S., Malgaroli, A. & Lamanna, J. 2023. Neural correlates of delay discounting in the light of brain imaging and non-invasive brain stimulation: What we know and what is missed. Brain Sciences, 13, 403.

Musholt, K. 2013. A philosophical perspective on the relation between cortical midline structures and the self. Frontiers in Human Neuroscience, 7, 536.

Nagy, K., Greenlee, M. W. & KovÁcs, G. 2012. The lateral occipital cortex in the face perception network: an effective connectivity study. Frontiers in psychology, 3, 141.

NIDA. 2020. What is drug addiction? [Online]. Available: https://nida.nih.gov/publications/drugs-brains-behavior-science-addiction/drug-misuse-addiction#ref [Accessed].

Northoff, G. 2017. Personal identity and cortical midline structure (CMS): do temporal features of CMS neural activity transform into “self-continuity”? Psychological Inquiry, 28, 122–131.

Oberlin, B. G., Albrecht, D. S., Herring, C. M., Walters, J. W., Hile, K. L., Kareken, D. A. & Yoder, K. K. 2015. Monetary discounting and ventral striatal dopamine receptor availability in nontreatment-seeking alcoholics and social drinkers. Psychopharmacology (Berl).

Oberlin, B. G., Shen, Y. I. & Kareken, D. A. 2020. Alcohol Use Disorder Interventions Targeting Brain Sites for Both Conditioned Reward and Delayed Gratification. Neurotherapeutics, 17, 70–86.

Ogilvie, D. M. 1987. The undesired self: A neglected variable in personality research. Journal of personality and social psychology, 52, 379.

Olszowy, W., Aston, J., Rua, C. & Williams, G. B. 2019. Accurate autocorrelation modeling substantially improves fMRI reliability. Nat Commun, 10, 1220.

Paternoster, R. & Bushway, S. 2009. Desistance and the” feared self”: Toward an identity theory of criminal desistance. The journal of criminal law and criminology, 1103–1156.

Petry, N. M., Bickel, W. K. & Arnett, M. 1998. Shortened time horizons and insensitivity to future consequences in heroin addicts. Addiction, 93, 729–738.

Pruim, R. H. R., Mennes, M., Van Rooij, D., Llera, A., Buitelaar, J. K. & Beckmann, C. F. 2015. ICA-AROMA: a robust ICA-based strategy for removing motion artifacts from fMRI data. Neuroimage, 112, 267–277.

Qin, P. & Northoff, G. 2011. How is our self related to midline regions and the default-mode network? Neuroimage, 57, 1221–33.

Ritchie, J. B., Montesinos, S. & Carter, M. J. 2024. What is a visual stream? Journal of Cognitive Neuroscience, 36, 2627–2638.

Rust, N. C. & Dicarlo, J. J. 2010. Selectivity and tolerance (“invariance”) both increase as visual information propagates from cortical area V4 to IT. Journal of Neuroscience, 30, 12978–12995.

Schacter, D. L., Addis, D. R. & Buckner, R. L. 2007. Remembering the past to imagine the future: the prospective brain. Nat Rev Neurosci, 8, 657–61.

Şenel, G. & Slater, M. Conversation with your future self about nicotine dependence. International Conference on Virtual Reality and Augmented Reality, 2020. Springer, 216–223.

Seo, D., Lacadie, C. M., Tuit, K., Hong, K.-I., Constable, R. T. & Sinha, R. 2013. Disrupted ventromedial prefrontal function, alcohol craving, and subsequent relapse risk. JAMA psychiatry, 70.

Shen, Y. I., Nelson, A. J. & Oberlin, B. G. 2022. Virtual reality intervention effects on future self-continuity and delayed reward preference in substance use disorder recovery: pilot study results. Discover Mental Health, 2, 19.

Sheridan, T. B. 1992. Musings on telepresence and virtual presence. Presence: Teleoperators and Virtual Environments, 1, 120–126.

Sobell, M. B., Sobell, L. C., Klajner, F., Pavan, D. & Basian, E. 1986. The reliability of a timeline method for assessing normal drinker college students’ recent drinking history: utility for alcohol research. Addict Behav, 11, 149–61.

Spreng, R. N. 2012. The fallacy of a “task-negative” network. Frontiers in psychology, 3, 145.

Spreng, R. N., Gerlach, K. D., Turner, G. R. & Schacter, D. L. 2015. Autobiographical planning and the brain: Activation and its modulation by qualitative features. Journal of cognitive neuroscience, 27, 2147–2157.

Spreng, R. N. & Grady, C. L. 2010. Patterns of brain activity supporting autobiographical memory, prospection, and theory of mind, and their relationship to the default mode network. Journal of cognitive neuroscience, 22, 1112–1123.

Spreng, R. N., Stevens, W. D., Chamberlain, J. P., Gilmore, A. W. & Schacter, D. L. 2010. Default network activity, coupled with the frontoparietal control network, supports goal-directed cognition. Neuroimage, 53, 303–317.

Stawarczyk, D. & D’Argembeau, A. 2015. Neural correlates of personal goal processing during episodic future thinking and mind-wandering: An ALE meta-analysis. Hum Brain Mapp, 36, 2928–47.

Szpunar, K. K., Spreng, R. N. & Schacter, D. L. 2014. A taxonomy of prospection: Introducing an organizational framework for future-oriented cognition. Proceedings of the National Academy of Sciences, 111, 18414–18421.

Tomasi, D., Wang, R., Wang, G.-J. & Volkow, N. D. 2014. Functional connectivity and brain activation: a synergistic approach. Cerebral cortex, 24, 2619–2629.

Tracy, E. M., Laudet, A. B., Min, M. O., Kim, H., Brown, S., Jun, M. K. & Singer, L. 2012. Prospective patterns and correlates of quality of life among women in substance abuse treatment. Drug and alcohol dependence, 124, 242–249.

Trope, Y. & Liberman, N. 2010. Construal-level theory of psychological distance. Psychological review, 117, 440.

Van Gelder, J. L., Luciano, E. C., Weulen Kranenbarg, M. & Hershfield, H. E. 2015. Friends with my future self: Longitudinal vividness intervention reduces delinquency. Criminology, 53, 158–179.

Wang, A., Elman, I., Lowen, S., Blady, S., Lynch, K., Hyatt, J., O’Brien, C. & Langleben, D. 2015. Neural correlates of adherence to extended-release naltrexone pharmacotherapy in heroin dependence. Translational psychiatry, 5, e531–e531.

Weiss, R. D., Potter, J. S., Fiellin, D. A., Byrne, M., Connery, H. S., Dickinson, W., Gardin, J., Griffin, M. L., Gourevitch, M. N., Haller, D. L., Hasson, A. L., Huang, Z., Jacobs, P., Kosinski, A. S., Lindblad, R., Mccance-Katz, E. F., Provost, S. E., Selzer, J., Somoza, E. C., Sonne, S. C. & Ling, W. 2011. Adjunctive counseling during brief and extended buprenorphine-naloxone treatment for prescription opioid dependence: a 2-phase randomized controlled trial. Arch Gen Psychiatry, 68, 1238–46.

Wesley, M. J. & Bickel, W. K. 2014. Remember the future II: meta-analyses and functional overlap of working memory and delay discounting. Biol Psychiatry, 75, 435–48.

Whiteside, S. P. & Lynam, D. R. 2001. The five factor model and impulsivity: using a structural model of personality to understand impulsivity. Person. individ. Diff., 30, 669–689.

Wilcox, C. E., Abbott, C. C. & Calhoun, V. D. 2019. Alterations in resting-state functional connectivity in substance use disorders and treatment implications. Progress in Neuro-Psychopharmacology and Biological Psychiatry, 91, 79–93.

Yee, N. 2007. The Proteus effect: Behavioral modification via transformations of digital self-representation. Human communication research, 33, 271–290.

Yeo, B. T., Krienen, F. M., Sepulcre, J., Sabuncu, M. R., Lashkari, D., Hollinshead, M., Roffman, J. L., Smoller, J. W., Zollei, L., Polimeni, J. R., Fischl, B., Liu, H. & Buckner, R. L. 2011. The organization of the human cerebral cortex estimated by intrinsic functional connectivity. J Neurophysiol, 106, 1125–65.

